# The natural recovery of visuospatial neglect: a systematic review and meta-analysis

**DOI:** 10.1101/2024.02.05.24302248

**Authors:** Margot Juliëtte Overman, Elena Binns, Elise T Milosevich, Nele Demeyere

## Abstract

**Background:** Visuospatial neglect is a common consequence of stroke and is characterised by impaired attention to contralesional space. Currently, the extent and time course of recovery from neglect are not clearly established. This systematic review and meta-analysis aimed to determine the natural recovery trajectory of post-stroke neglect.

**Methods:** PsycInfo, Embase, and MEDLINE were searched for articles reporting recovery rates of neglect after stroke. Time since stroke was categorised into early (0-3 months), mid (3-6 months), or late (>6 months) recovery phases. Random-effects models for pooled prevalence were generated for each phase, and potential sources of heterogeneity were explored with meta-regressions. Methodological quality of each study was assessed using the Joanna Briggs Institute checklist, with low-quality studies excluded in sensitivity analyses.

**Results:** A total of 27 studies reporting data from 839 stroke survivors with neglect were included. Meta-analyses indicated a recovery rate of 42% in the early phase, which increased to 53% in the mid-recovery phase. Additional recovery in the late phase was minimal, with an estimated 56% recovery rate. Estimates were robust to sensitivity analyses. Meta-regressions showed significantly greater recovery in studies which included patients with left-hemisphere lesions (*ß*=0.275, *p*<0.05).

**Conclusions:** Most natural recovery from neglect occurs in the first three months, although additional gains can be expected up to 6 months post-stroke. Whilst a large proportion of patients recover from neglect, over 40% show persistent symptoms. Further research is needed on effective rehabilitation interventions, particularly focusing on patients most at risk of chronic visuospatial neglect.

## BACKGROUND

Visuospatial neglect is a common post-stroke syndrome characterised by inattention to stimuli in contralesional space, which cannot be attributed to primary sensory or motor deficits[1]. Neglect is a heterogeneous disorder, encompassing various subtypes which can occur in isolation or in conjunction [2]. Among others, neglect can affect distinct spatial regions (e.g. personal, peripersonal, and/or extrapersonal space) and manifest as person-centred (egocentric) or stimulus-centred (allocentric) spatial attention deficits[3]. Recent estimates indicate that ∼30% of stroke patients present with neglect acutely post-stroke, with prevalence rates typically being higher after right-than left-hemisphere lesions[4]. Neglect negatively impacts a wide range of outcomes, including discharge destination and independence in daily activities[5–7], rehabilitation efficacy[8], and quality of life[9,10]. An improved understanding of the persistence of neglect over time is therefore of high clinical relevance. However, there is currently no consensus regarding either the extent or time course of natural recovery from neglect.

Natural or spontaneous recovery refers to the improvement of function determined by the progression of time[11]. The degree of natural recovery after stroke varies substantially across domains and has been most extensively studied in the context of motor impairment, although several overarching principles have been identified. First, significant recovery typically occurs within three months post-stroke[12]. Second, recovery tends to be proportional to the severity of acute deficits[13]. Patients with more severe impairments are expected to make significant improvements over time, but may be less likely to reach formal recovery thresholds compared to those with milder deficits. Third, functional gains beyond the first three months are more common for cognitive difficulties than motor impairments[12], with residual improvements observed years after stroke for language, working memory, and global cognition[14–16].

The specific trajectory of visuospatial neglect recovery, however, remains underdefined. Therefore, the primary aim of this systematic review is to identify the longitudinal pattern of natural recovery from visuospatial neglect after stroke. Importantly, stroke survivors who do not meet criteria for ‘full recovery’ may still demonstrate clinically significant gains in function over time. Accordingly, the secondary aim is to examine natural *improvement* of neglect over time. Finally, previous research suggests certain patient and study characteristics can impact on the observed recovery of neglect. Specifically, lesion side, severity of stroke and neglect, and time of the first assessment may influence reported recovery rates[17–19]. We hypothesise that (a) left-hemisphere lesions are associated with higher recovery rates than right-hemisphere stroke[19,20], (b) greater severity of stroke or neglect at baseline is associated with reduced likelihood of recovery[10], and (c) studies which conducted initial assessments of neglect early post-stroke (days) will report higher levels of recovery than those recruiting patients at a later time, as acute neglect can resolve within days after stroke[19,20].

## METHODS

The protocol for this systematic review was registered with PROSPERO (CRD42023388763). The review was reported according to Preferred Reporting Items for Systematic Reviews and Meta-analysis (PRISMA) guidelines[21].

### Search strategy

The OVID platform was used to search PsycInfo, Embase, and MEDLINE databases from inception to 13 December 2022. The search strategy was developed in consultation with a librarian and a neuropsychologist specialising in stroke. Title, abstract, and relevant topic terms were searched with Boolean operators using the keywords ‘stroke’, ‘neglect’, and ‘neuropsychological assessment’ (see Supplemental Material for the detailed search strategy). References in the selected journal papers were reviewed to identify additional relevant studies.

### Eligibility criteria

Study inclusion criteria were: (1) peer-reviewed observational studies, (2) published in English language, (3) included patients who developed visuospatial neglect following stroke, (4) assessed neglect with a standardised test, and (5) included ≥2 different time points. Case studies, commentaries, review articles, and conference abstracts were not considered. Articles which included patients below the age of 18, patients with dementia, or patients who had neglect prior to a stroke diagnosis were excluded. Studies were also excluded if they involved an intervention or treatment element, or if only tests for global cognition were used.

### Screening and data extraction

Following the removal of duplicates, screening of titles, abstracts and full texts was conducted by E.B. The resulting list of studies was reviewed by a second author (M.J.O.), with any disagreements regarding study eligibility being reconciled in consultation with a third reviewer (N.D.). Data from each eligible report were independently extracted by two authors (E.B. and M.J.O.) using a predefined extraction template, with any discrepancies resolved through discussion with all authors. Data extracted included authors, publication year, sample size, number of patients with neglect, country, study setting, age, sex, stroke aetiology, stroke severity, thrombolysis or thrombectomy treatment, standard rehabilitation procedures, neuropsychological assessments used, average performance on neglect assessments, and time of assessment.

### Outcomes

The primary outcome of interest was the proportion of stroke survivors with neglect at each time point. For every study, only patients who were diagnosed with neglect at baseline and completed follow-up were included in analyses. It was not possible to stratify studies based on neglect characteristics (e.g. egocentric vs. allocentric) due to limited reporting of neglect subtypes. Where studies reported recovery rates for different neglect subtypes but did not specify which patients presented with co-occurring symptoms, we selected the subtype with the largest sample size for analyses. Recovery percentage was calculated at every assessment: (patients recovered from neglect/total number of patients) x 100.

Across all studies, scores on the Behavioural Inattention Task (BIT)[22] were most frequently reported. The conventional BIT consists of six subtests, including line crossing, letter cancellation, star cancellation, figure and shape copying, line bisection, and representational drawing tasks. The maximum total score on the BIT is 146, with scores below 129 being indicative of visuospatial neglect[23]. To assess improvement of neglect severity over time as a secondary outcome, average scores on the BIT were extracted from each study where available.

### Quality assessment

The methodological quality of included articles was assessed using the Joanna Briggs Institute (JBI) critical appraisal checklist for studies reporting prevalence data[24]. Study quality was determined by the number of items with a ‘yes’ response (max. 9), with the total score converted into a percentage. Studies scoring <50% were categorised as low-quality, 50-69% as moderate quality, and ≥70% as high-quality[25]. All studies were rated independently by two authors (E.B. and M.J.O.), with any differences in ratings resolved through discussion with all authors.

### Statistical analysis

All analyses were carried out using R software (version 4.1.2). For each study, time since stroke was categorised into early (0-3 months), mid (3-6 months), and late (>6 months) recovery phases. Meta-analyses of the key outcomes of interest were stratified by recovery phase. Where patients were assessed multiple times within the same phase, the time point with the largest sample size was used for meta-analyses. Pooled estimates were generated with random-effects meta-analyses using the ‘meta’ package. To estimate overall proportion of neglect recovery, the ‘metaprop’ function was applied using the inverse variance method with a Freeman-Tukey Double Arcsine transformation. The level of heterogeneity was estimated using the restricted maximum likelihood method, with significance indicated by Cochran Q test *p*-values <0.05. Heterogeneity between studies was quantified by the resulting *I*^2^ statistic and interpreted as low (<25%), medium (50-75%), or high (>75%)[26]. All results are presented as forest plots and associated 95% confidence intervals (CIs). In sensitivity analyses, studies rated as low-quality on the JBI were excluded to evaluate the robustness of results. Where sufficient data were available, meta-regressions were performed to explore variability in recovery outcomes. Key predictors of interest were lesion side, stroke and neglect severity, and time of first assessment (≤7 days versus >7 days post-stroke).

### Data availability

The full R code and extracted data can be freely accessed through https://osf.io/zwkty/.

## RESULTS

### Study selection

The initial search identified a total of 4,130 records. Following removal of duplicates, the titles and abstracts of 2,321 publications were screened. The full texts of 131 articles were reviewed, with 36 studies meeting the inclusion criteria. Nine papers did not clearly identify which patients with neglect versus non-neglect at baseline completed follow-up assessments and were therefore excluded, resulting in a total of 27 publications. The study selection process is displayed in Figure 1.

**Figure 1.**
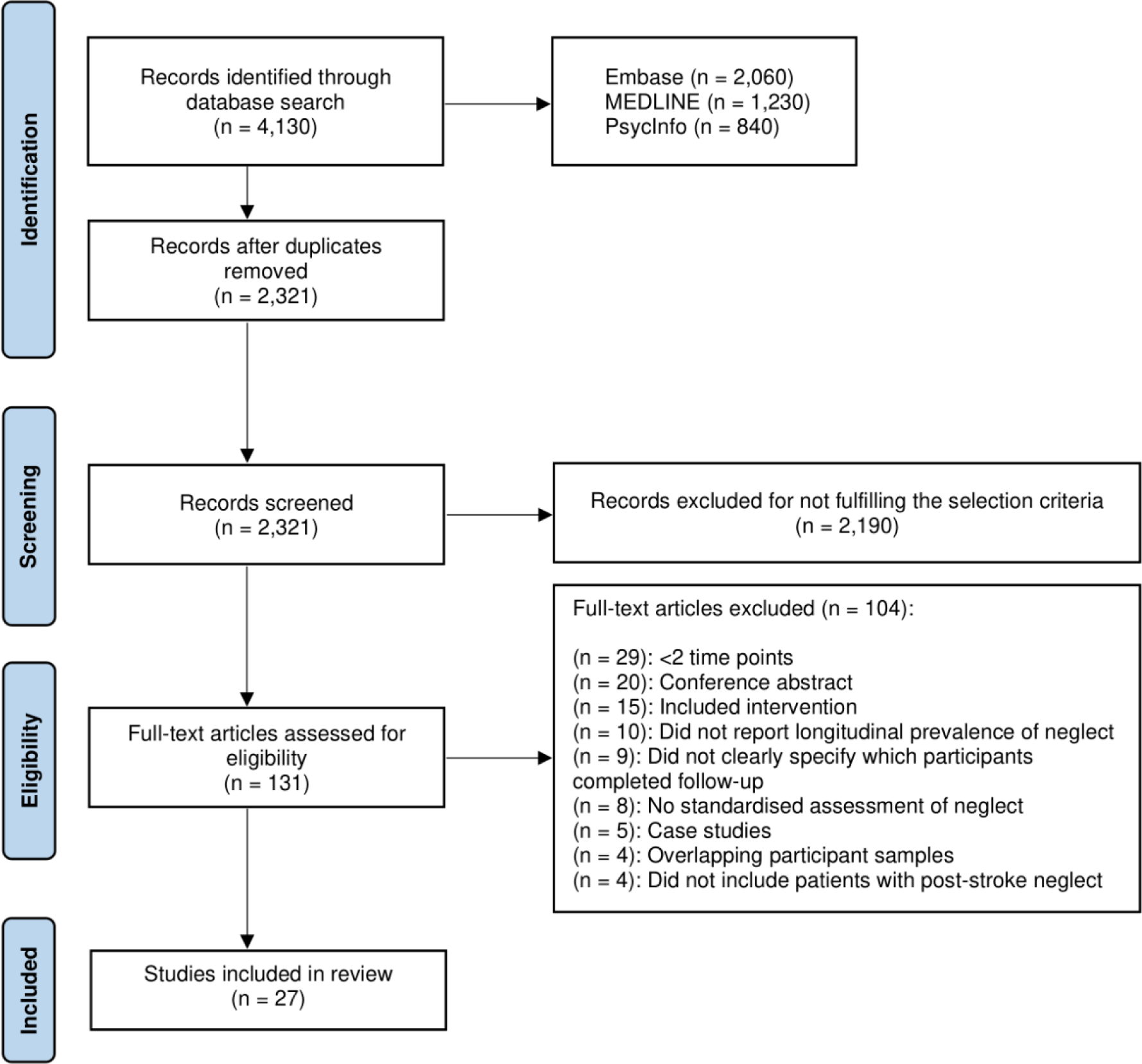
PRISMA flow chart of study selection

### Population and study characteristics

Table 1 provides a summary of the characteristics of all studies included in the meta-analyses. The 27 included studies reported data from a total of 839 stroke survivors with neglect at baseline. Sample sizes ranged from 6-142 patients (median 23 patients). Stroke survivors were predominantly recruited from hospital sites including rehabilitation centres (n=22), while 1 study recruited from a regional stroke register and 4 studies did not explicitly report recruitment setting. Studies were conducted in Europe (n=21), Australia (n=2), Asia (n=2), and North America (n = 2).

**Table 1.**
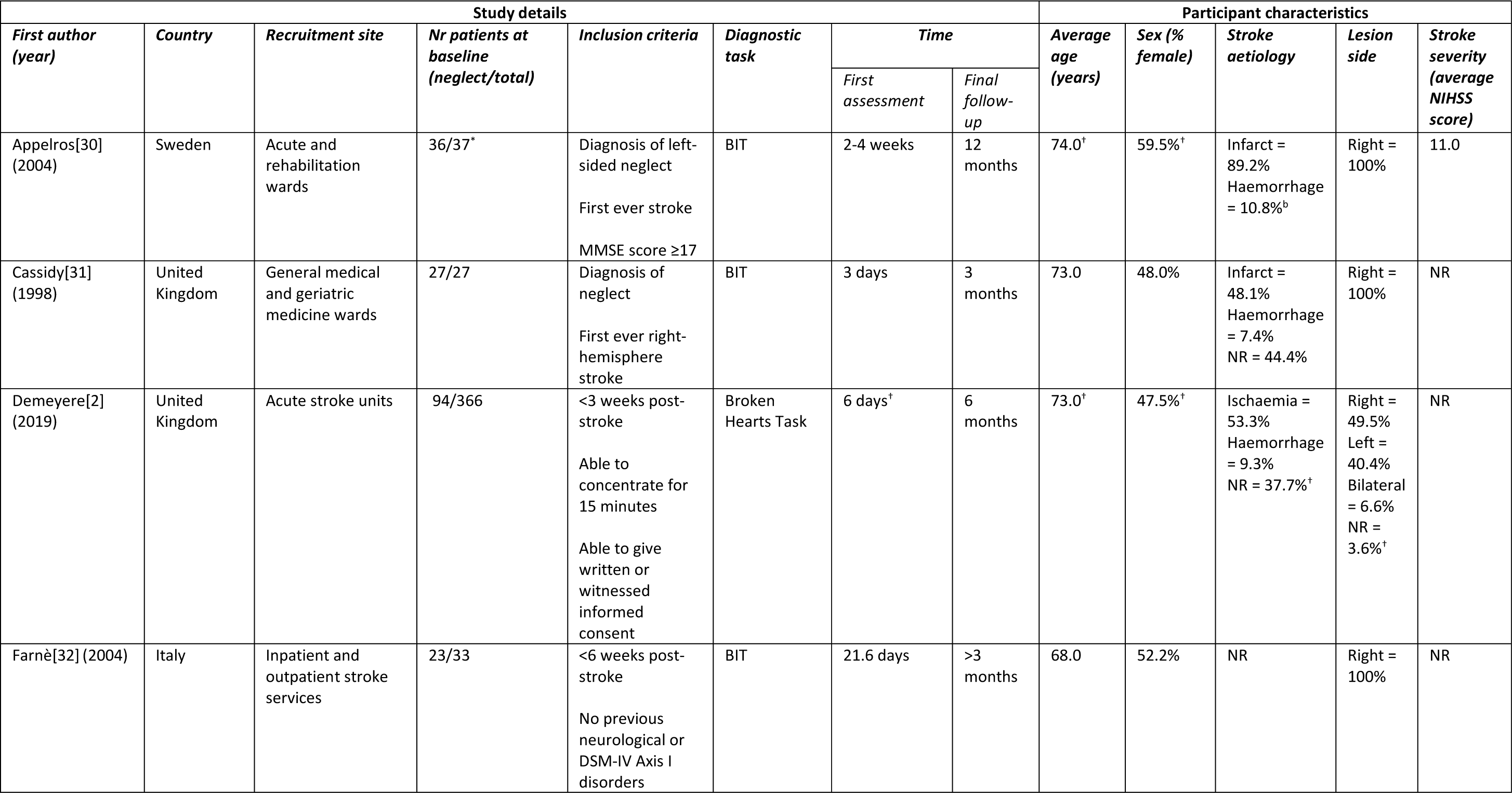
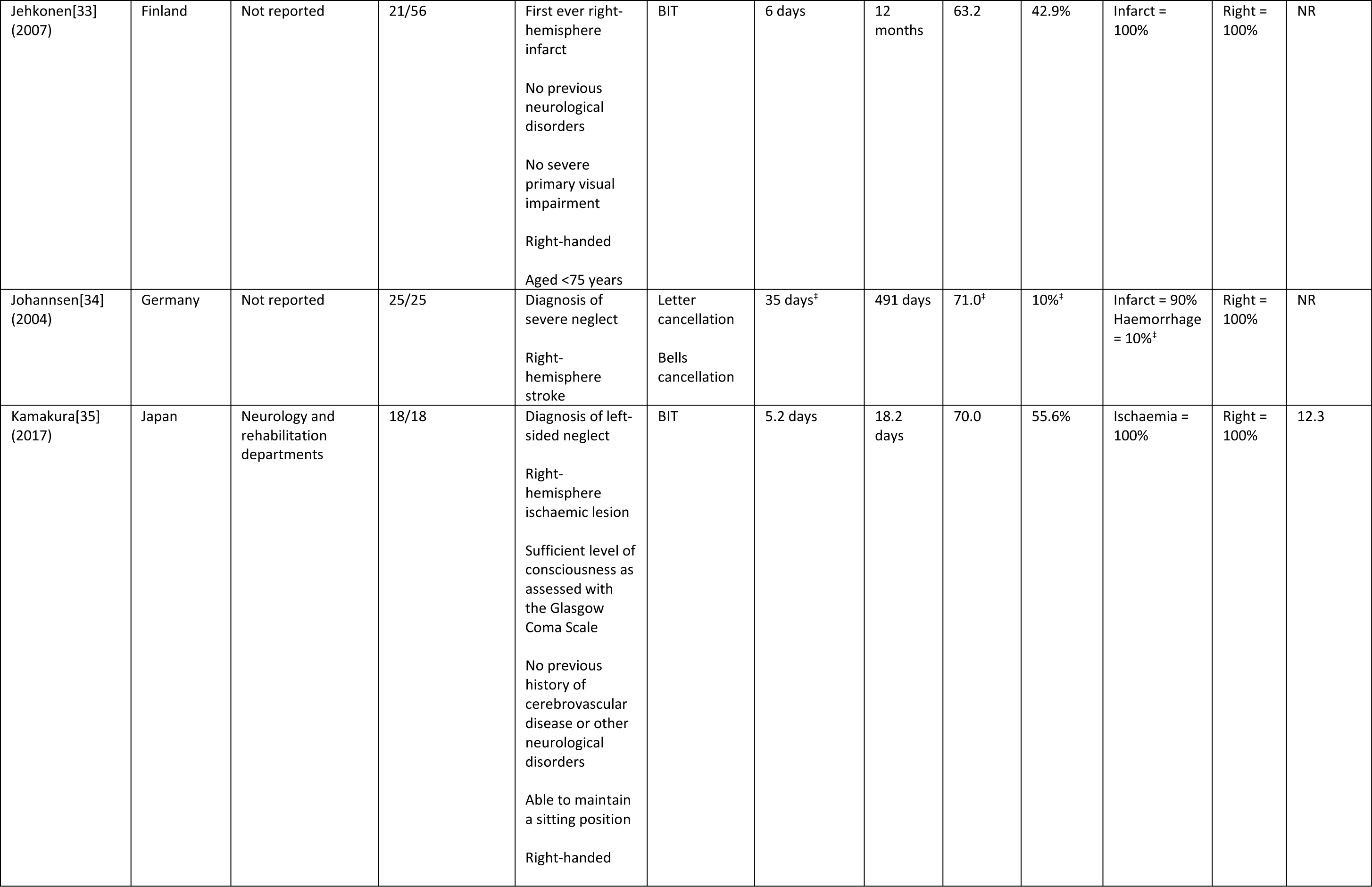
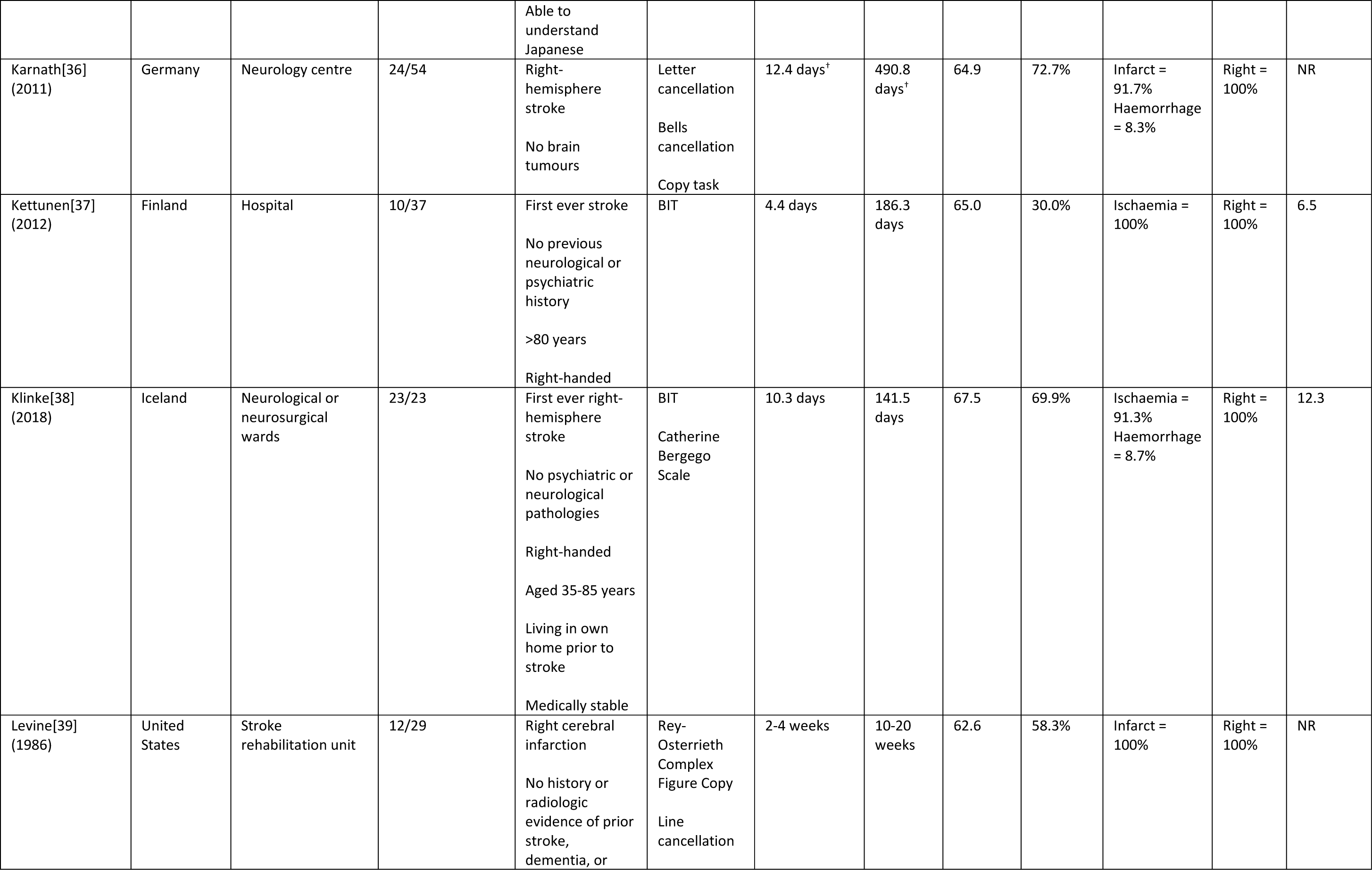
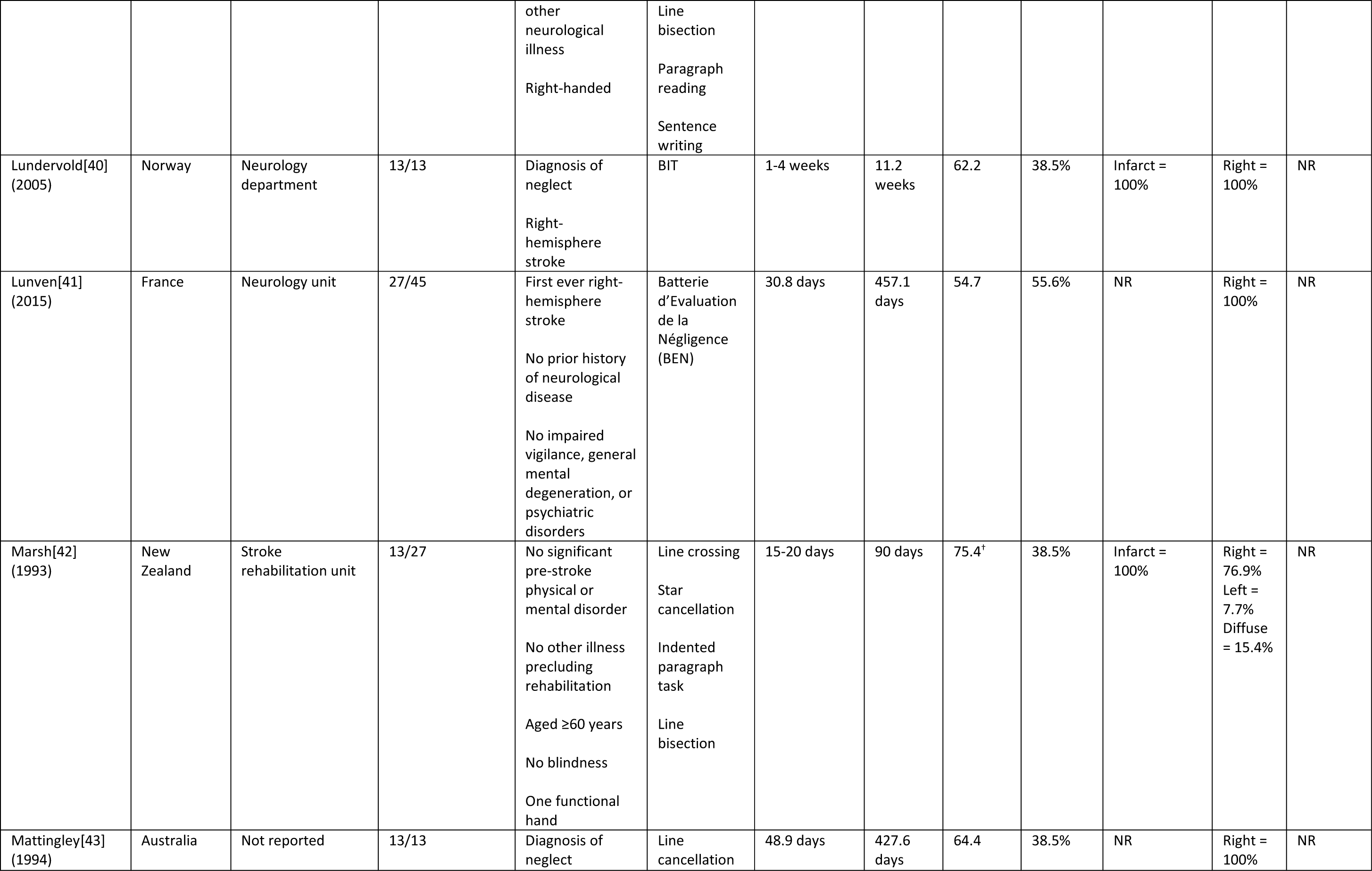
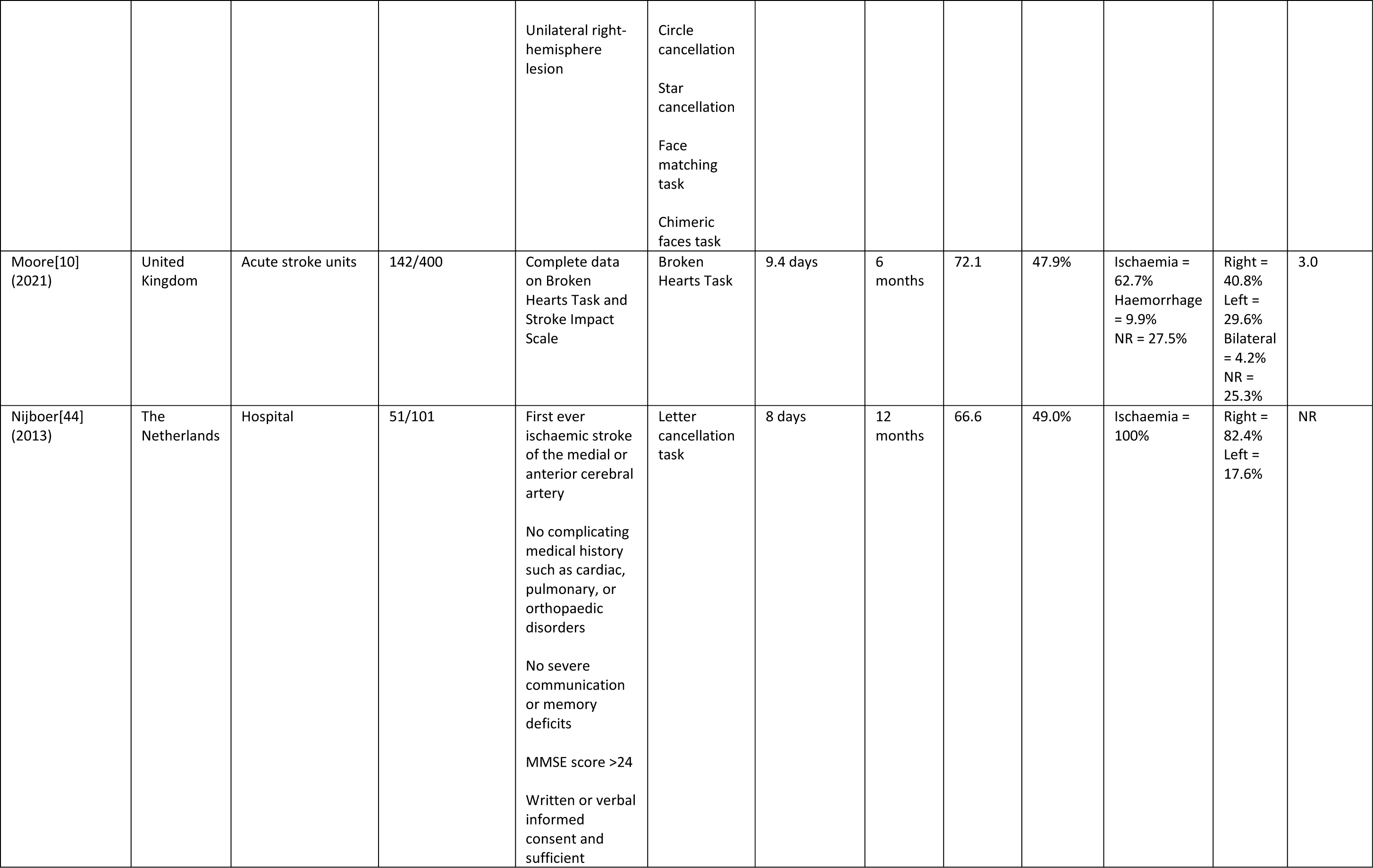
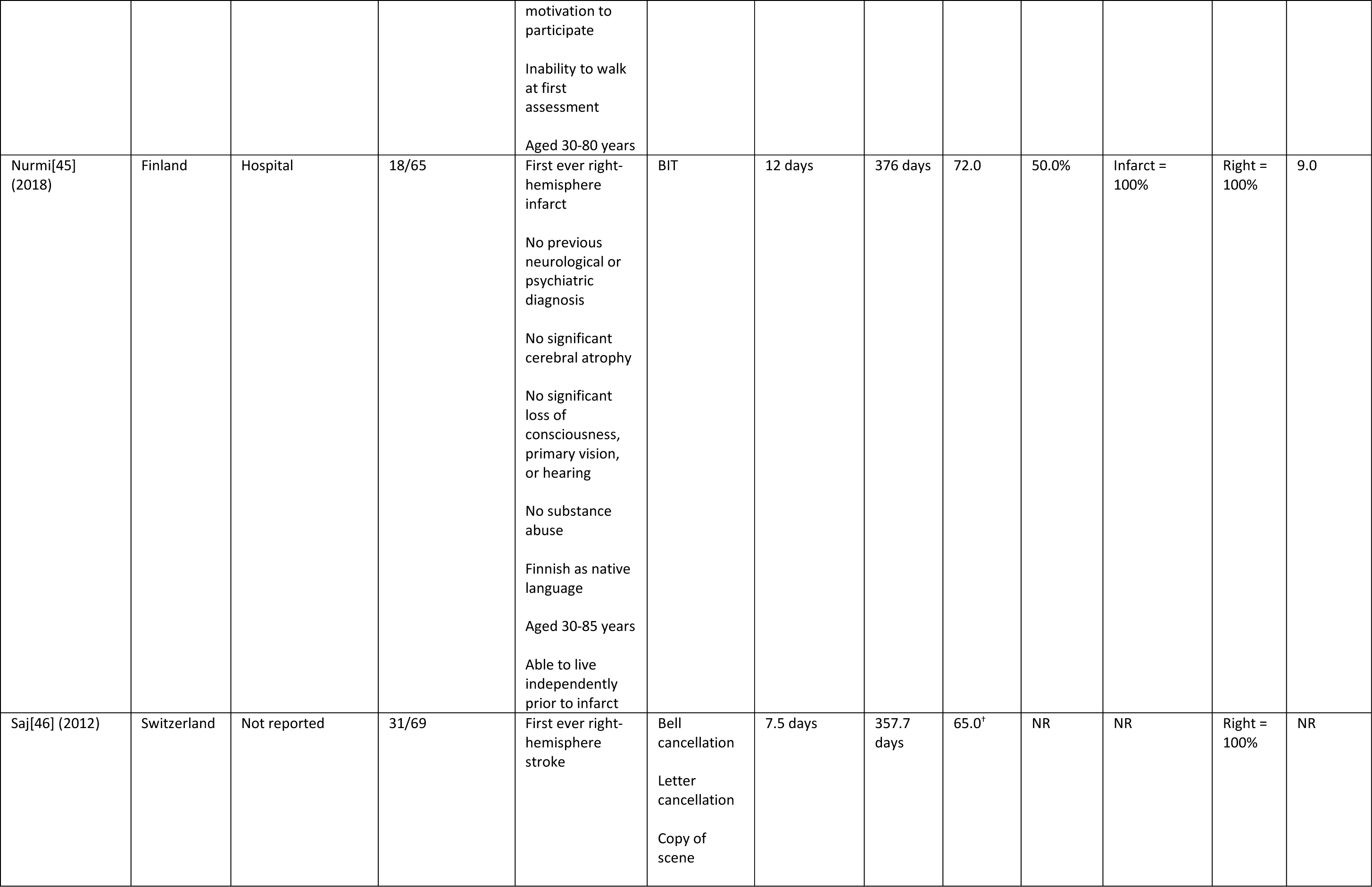
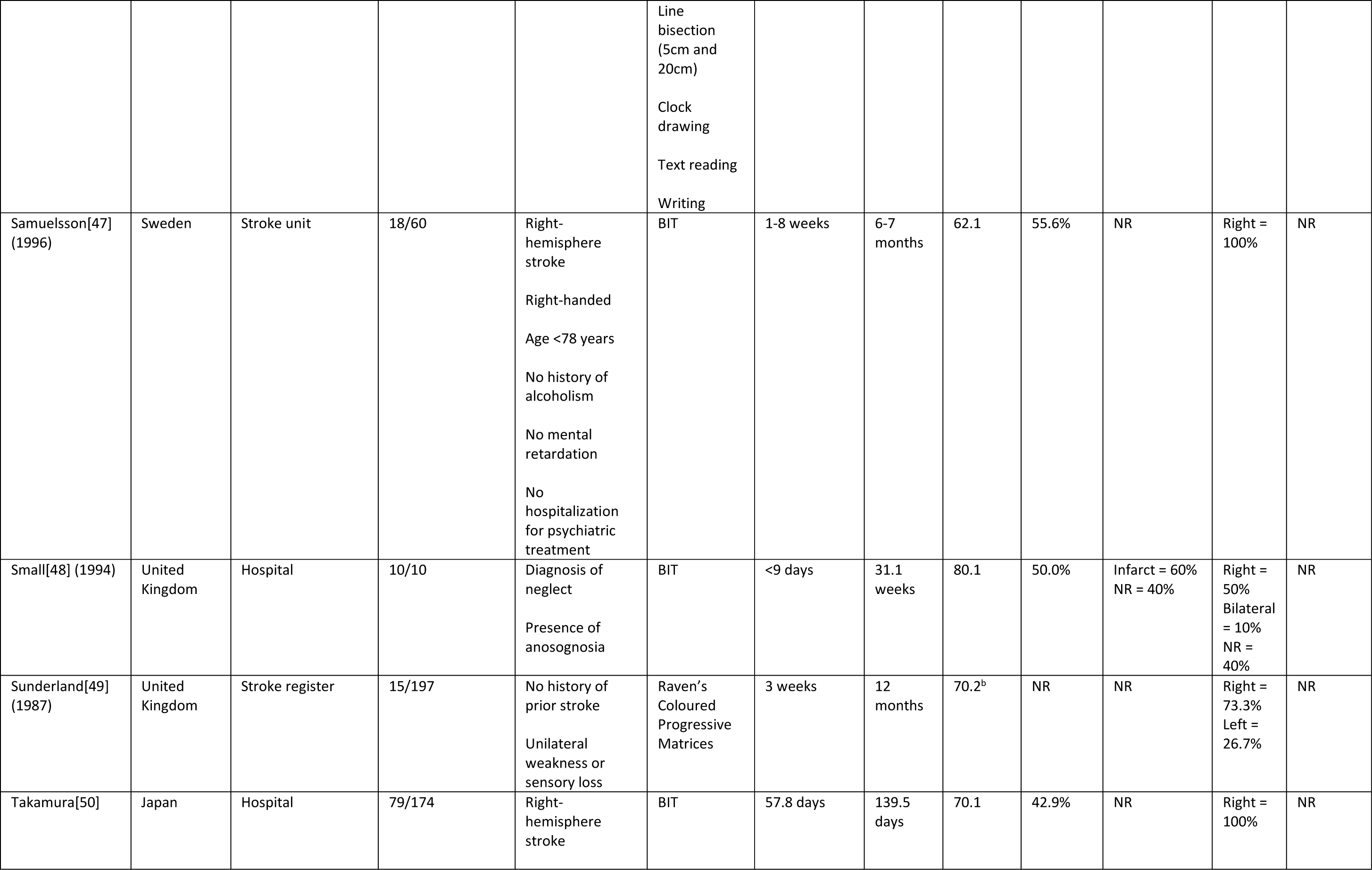
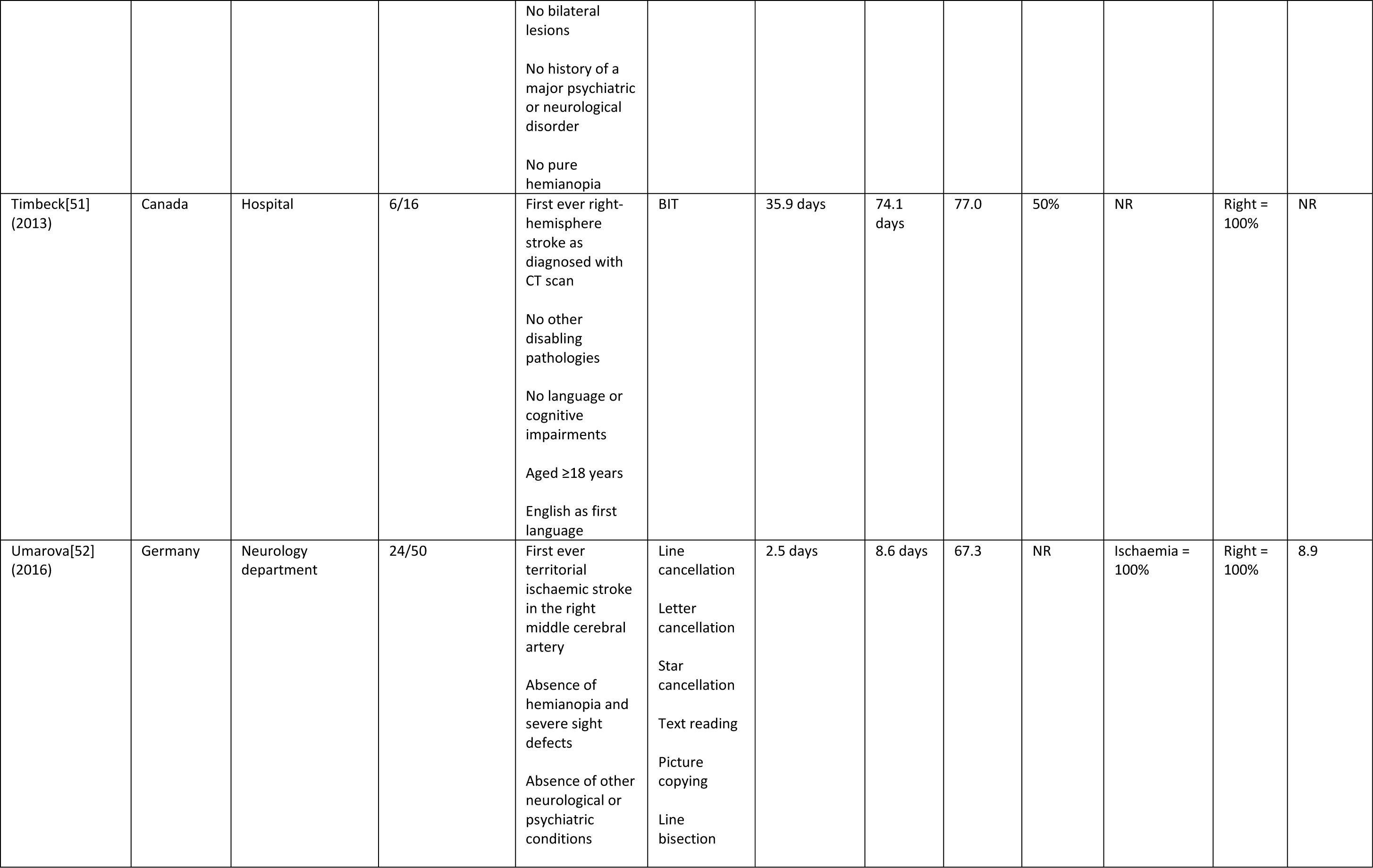
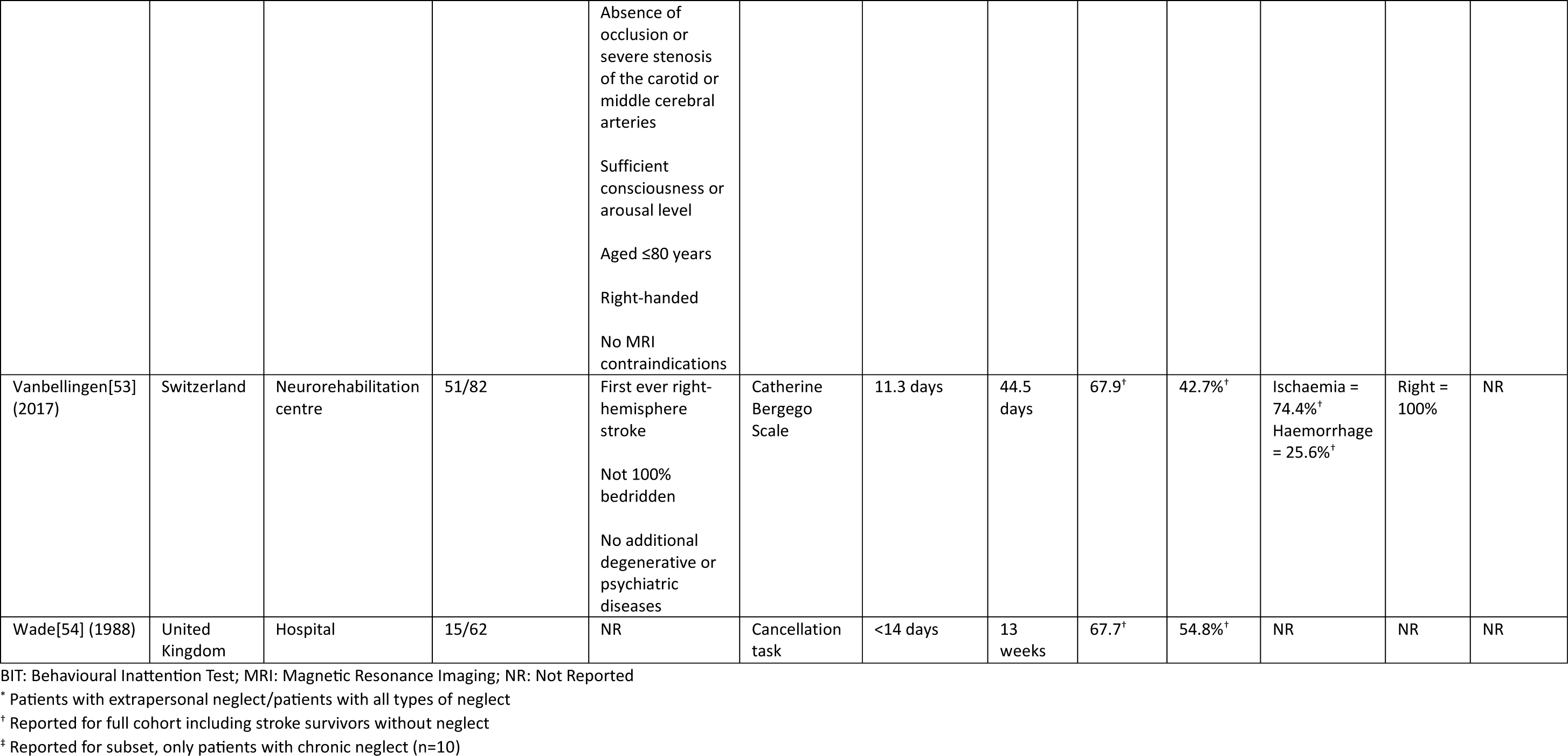
Summary of study and patient characteristics.

The average age of stroke survivors with neglect was reported in 19 studies and ranged from 54.7-80.1 years. Ischaemic stroke was most common (n=331 (39.5%) patients), followed by haemorrhagic stroke (n=20 (2.4%) patients). Aetiology was unreported or unknown for 488 (58.2%) patients. Twenty studies only included patients with a right-hemisphere lesion. Overall, 600 (71.5%) patients had a right-hemisphere stroke, 56 (6.7%) patients had a left-hemisphere stroke, 9 (1.1%) patients had a diffuse or bilateral stroke, and lesion side was unknown for 174 (20.7%) patients. Baseline assessment ranged from 2.5-48.9 days post-stroke, and total follow-up time ranged from 8.6-491 days post-stroke.

A diagnosis of visuospatial neglect was most often based on BIT scores (n=13), with the presence of neglect being determined by total scores <129 and/or performance below cut-off scores for ≥2 of 6 subtasks. The remaining studies used (a combination of) cancellation tasks (n=11), figure copying (n=4), text reading (n=4), line bisection (n=2), writing (n=2), the Catherine Bergego Scale[27] (n=2), face matching tasks (n=1), Raven’s Coloured Progressive Matrices[28] (n=1), or a full neglect test battery (n=1). Stroke severity as measured with the National Institutes of Health Stroke Scale (NIHSS) was reported in 7 studies, with average scores ranging from 3-12.3 (i.e., mild to moderately severe[29]).

### Quality assessment

Thirteen studies were rated as high-quality [24], 8 studies rated as moderate quality, and 6 studies as low-quality. All studies measured neglect with standard and reliable methods, and all but one study used validated measures for the identification of neglect. Only 4 studies had a sufficient sample size, highlighting the need for large-scale studies of visuospatial neglect. Additionally, sample frame proved to be a problematic item with only 6 studies representing the target population. The main reason for failing this criterion was the exclusion of patients with a left-hemisphere stroke. Visuospatial neglect has consistently been observed after left-hemisphere lesions[55], and exclusion of left-hemisphere stroke patients was therefore treated as non-representative of the entire target population[56]. An item-by-item overview of the quality assessment for all studies is presented in Table 2.

**Table 2.** Quality assessment of included studies.

| **Study** | **Q1**<br>*Sample frame* | **Q2**<br>*Sampling method* | **Q3**<br>*Sample size* | **Q4**<br>*Study description* | **Q5**<br>*Sample coverage* | **Q6**<br>*Validity of assessment* | **Q7**<br>*Standardised measurement* | **Q8**<br>*Statistical analysis* | **Q9**<br>*Response rate* | **Total score** | **Quality rating** |
| --- | --- | --- | --- | --- | --- | --- | --- | --- | --- | --- | --- |
| Appelros[30] | N | Y | N | Y | Y | Y | Y | Y | U | 6 | moderate |
| Cassidy[31] | U | Y | N | Y | Y | Y | Y | Y | Y | 7 | high |
| Demeyere[2] | Y | Y | Y | Y | Y | Y | Y | Y | Y | 9 | high |
| Farné[32,33] | Y | U | N | Y | U | Y | Y | Y | U | 5 | moderate |
| Jehkonen[33] | N | Y | N | Y | N | Y | Y | Y | U | 5 | moderate |
| Johannsen[34] | U | Y | N | N | N | Y | Y | Y | Y | 5 | moderate |
| Kamakura[35] | N | Y | N | Y | Y | Y | Y | Y | N | 6 | moderate |
| Karnath[36] | N | Y | N | Y | Y | Y | Y | Y | Y | 7 | high |
| Kettunen[37] | N | Y | N | Y | Y | Y | Y | Y | Y | 7 | high |
| Klinke[38] | N | Y | N | Y | Y | Y | Y | Y | Y | 7 | high |
| Levine[39] | N | U | N | Y | Y | Y | Y | Y | N | 5 | moderate |
| Lundervold[40] | U | Y | N | N | U | Y | Y | Y | N | 4 | low |
| Lunven[41] | N | Y | N | Y | Y | Y | Y | Y | Y | 7 | high |
| Marsh[42] | Y | Y | N | Y | Y | Y | Y | Y | Y | 8 | high |
| Mattingley[43] | U | U | N | Y | Y | Y | Y | Y | U | 5 | moderate |
| Moore[10] | Y | Y | Y | Y | Y | Y | Y | Y | Y | 9 | high |
| Nijboer[44] | Y | U | Y | Y | Y | Y | Y | Y | Y | 8 | high |
| Nurmi[45] | N | Y | N | Y | Y | Y | Y | Y | Y | 7 | high |
| Saj[46] | N | U | N | N | U | Y | Y | Y | Y | 4 | low |
| Samuelsson[47] | N | Y | N | Y | Y | Y | Y | Y | Y | 7 | high |
| Small[48] | N | U | N | N | N | Y | Y | U | N | 2 | low |
| Sunderland[49] | Y | Y | N | N | N | N | Y | Y | Y | 5 | moderate |
| Takamura[50] | N | U | N | Y | N | Y | Y | Y | U | 4 | low |
| Timbeck[51] | N | Y | N | Y | Y | Y | Y | Y | Y | 7 | high |
| Umarova[52] | U | U | N | N | Y | Y | Y | Y | U | 4 | low |
| Vanbellingen[53] | N | Y | Y | Y | Y | Y | Y | Y | Y | 8 | high |
| Wade[54] | U | Y | N | N | U | Y | Y | U | N | 3 | low |

### Natural recovery of neglect

Figure 2 displays the reported natural recovery of neglect over time for all 27 studies. Meta-analyses were stratified by recovery phase (see Figure 3). In the early recovery phase (0-3 months), pooled data from 12 studies including 262 patients indicated an estimated recovery rate of 42% (CI=20-64%, *I*^2^=91%). The proportion of patients who recovered from neglect increased further in the mid-recovery phase (3-6 months) to 53% (CI=35-70%, *I*^2^=89%) based on 11 studies with a total of 426 patients. There was minimal further recovery in the late phase (>6 months), with an estimated recovery prevalence of 56% (CI=41-70%, *I*^2^=77%) across 12 studies with 257 patients. Results from sensitivity analyses, which excluded low-quality studies, were very similar to the main meta-analyses with estimated recovery rates of 38% (early), 53% (mid), and 56% (late) phases (see Supplemental Material, Figure S1).

**Figure 2.**
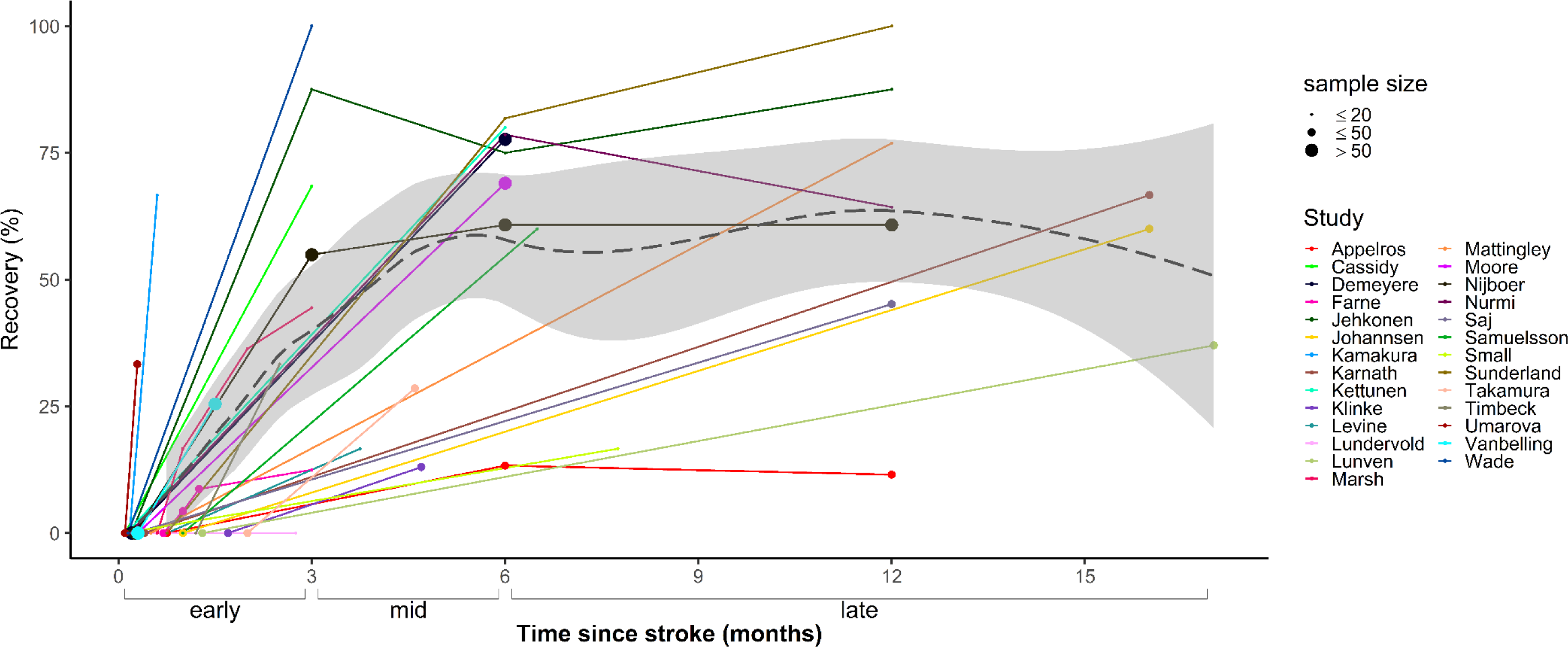
Natural recovery of visuospatial neglect in the early, mid, and late phases after stroke. Individual lines indicate the proportion of recovered patients at each assessment point for every study, with dot size reflecting sample size. The LOESS line (dashed) shows the estimated smooth fit of the regression model of recovery rates predicted by time across all studies, with the 95% confidence interval shaded in grey. Most recovery occurred in the early phase (0-3 months), with smaller increases observed in the mid-recovery phase (3-6 months). No additional recovery was observed in the late recovery phase (>6 months).

**Figure 3.**
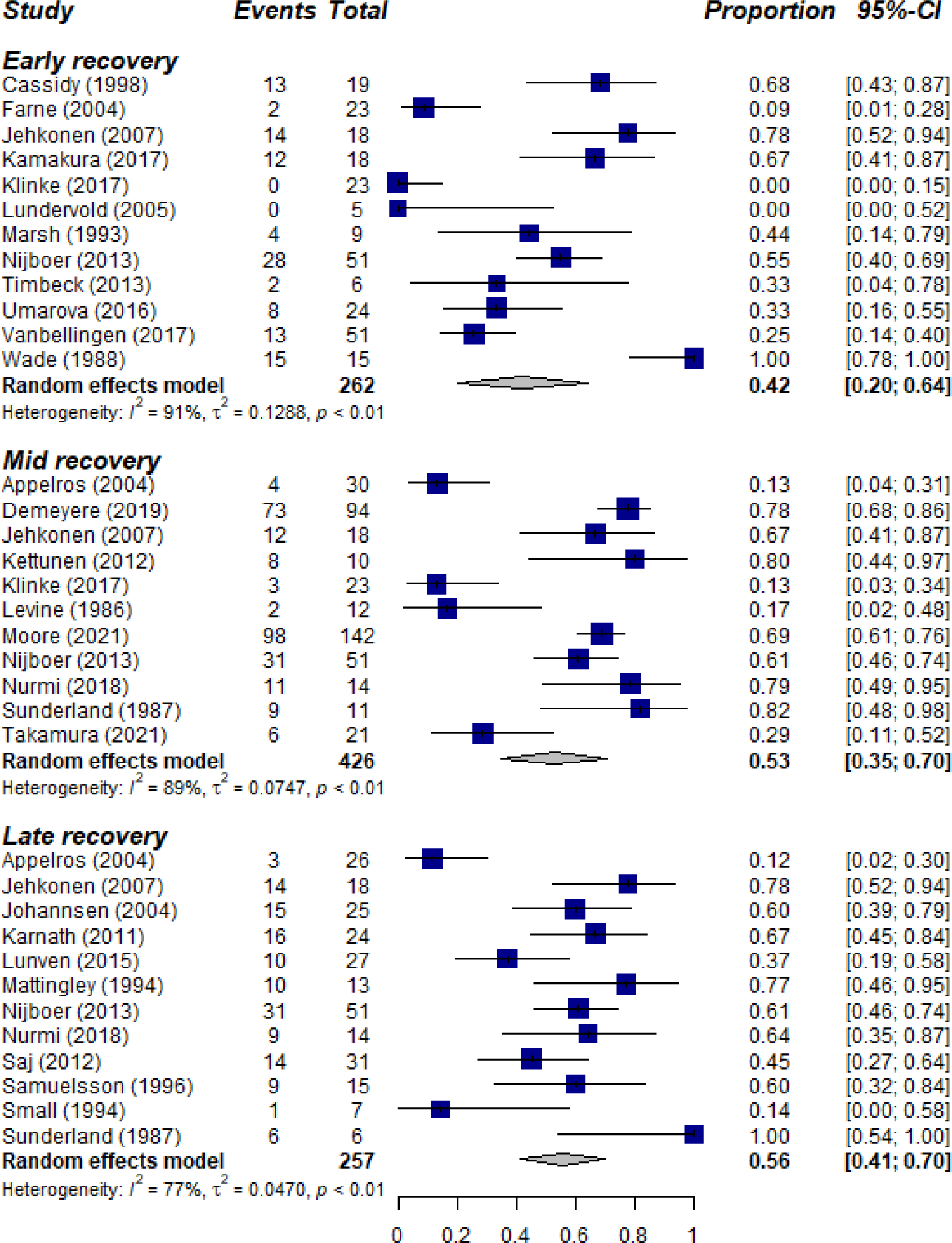
Forest plots of neglect recovery stratified by phase.

### Sources of heterogeneity

Heterogeneity of studies was high (*I*^2^ >75%) in all three phases of recovery. Using meta-regressions, we explored factors which were hypothesised to contribute to variable outcomes across studies. First, we assessed the impact of inclusion versus exclusion of patients with left-hemisphere lesions. We found that studies which included left-hemisphere stroke patients reported greater recovery rates compared with studies that only included right-hemisphere lesions (*ß*=0.275, *p*<0.05). We also examined whether timing of baseline assessment, which varied substantially between studies (see Figure 2), affected recovery rates. Results showed no evidence that time of first assessment moderated reported rates of recovery (*ß*=-0.211, *p*=0.101). Due to limited reporting of NIHSS scores and neglect severity, it was not possible to formally investigate the influence of stroke and neglect severity on recovery.

### Improvement of neglect

To determine improvement of neglect symptoms over time, changes in scores on the BIT[22] were examined. A total of 8 studies with 128 patients reported average or individual BIT scores at multiple time points (Figure 4). Due to the limited number of studies per phase, it was not possible to carry out meta-analyses using BIT scores. Therefore a descriptive overview of the BIT data is provided. At baseline, patients across studies had a mean score of 87.3 (mean range=56.3-121.7). In the early recovery phase, there was a substantial improvement in scores with an average performance of 122.1 (mean range =96.2-136.8). Scores increased further in the mid-recovery phase to a mean of 138.1 (mean range=114.1-139.9) but remained stable in the late recovery phase (M=138.0, mean range=137.9-138.2).

**Figure 4.**
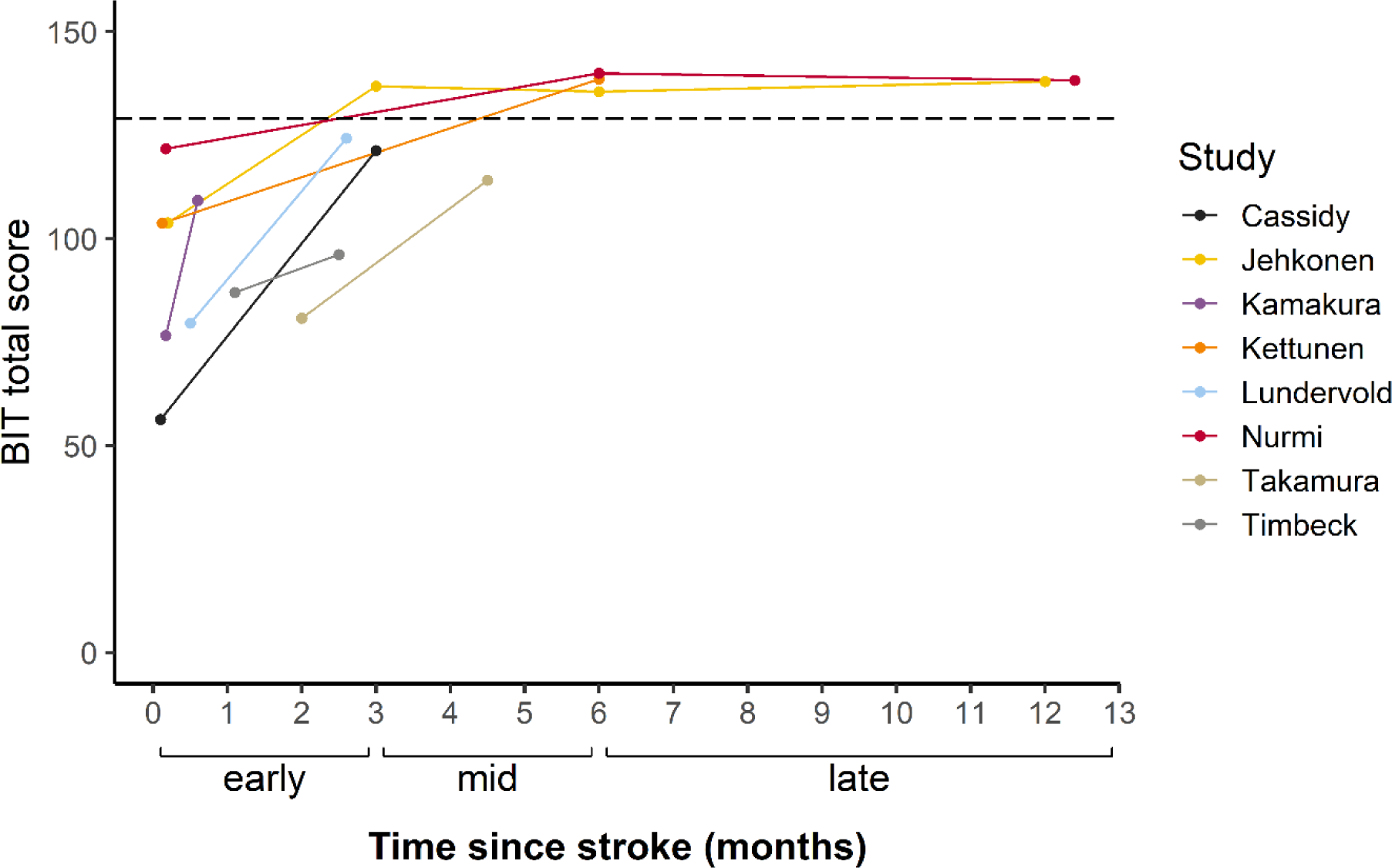
Changes in average BIT scores over time. Individual lines indicate mean scores reported by each study. The dashed line represents the cut-off score for a diagnosis of neglect.

## DISCUSSION

This systematic review and meta-analysis of 27 studies showed that 42% of stroke survivors with neglect recover within the first three months after stroke. The pooled prevalence of neglect recovery increased further between three to six months post-stroke, with an average of 53% of patients meeting recovery criteria. There was no evidence of clinically significant additional natural recovery beyond this period, with an estimated recovery rate of 56% between 6 to 17 months after stroke. To gain a more detailed understanding of changes in neglect severity over time, scores on a standardised assessments of neglect were investigated. In a subset of 8 studies, mean BIT scores were just below the cut-off for normal performance at three months post-stroke. This showed that although not all stroke survivors met the formal threshold of recovery, patients on average showed material improvements of neglect symptoms in the early recovery phase. In line with the main meta-analyses, additional but less extensive improvements were observed up to six months post-stroke, with minimal changes in longer-term follow-up assessments. Collectively, these findings are consistent with the literature on motor and cognitive impairments post-stroke, which indicates that most natural recovery occurs within the first three months[12,57]. However, our results demonstrate that patients with visuospatial neglect can expect smaller but clinically significant gains up to six months post-stroke.

### Variability in neglect recovery rates

There was substantial heterogeneity across studies, with some reporting complete recovery while others observed chronic neglect in all stroke survivors. There are likely several factors contributing to this variability in patient outcomes. First, as highlighted in our study quality assessment, sample sizes tended to be relatively small. Fourteen studies involved fewer than 20 participants, and only four studies had a sample size of 50 or more patients with neglect at baseline. Notably, three studies which had fewer than 10 participants at follow-up reported either minimal[40,48] or complete recovery[49]. Such extreme outcomes can likely be attributed, at least in part, to insufficient sample sizes. Additionally, the quality assessment indicated that most studies excluded patients with left-hemisphere lesions. Although neglect is more prevalent after right-hemisphere damage[58], a recent review showed that 20% of all stroke survivors with left-hemisphere lesions are also affected by neglect[4]. Moreover, the impact of neglect on rehabilitation outcomes is similar for left- and right-hemisphere stroke survivors[4,58]. It is therefore important to understand the recovery of neglect following lesions within either hemisphere. Most studies claim neglect is more persistent after right-than left-hemisphere stroke[19,20,59]. This review supports this notion as we observed greater recovery in studies which included left-hemisphere stroke patients. However, as few studies reported recovery outcomes separately for left- and right-hemisphere stroke patients, it was not possible to directly contrast recovery patterns between these two groups.

We also hypothesised that studies which conducted the baseline assessment of neglect within the first week after stroke would observe higher levels of recovery relative to studies which recruited patients at a later time. Contrary to our expectations, meta-regressions showed no evidence that the time at which the first assessment was completed influenced reported recovery rates. However, only 6 studies examined patients within the first week after stroke, with 2 studies completing neuropsychological examinations in the first three days. Given the limited amount of information on these initial days post-stroke, it is possible that this review underestimates potential rapid recovery from hyperacute neglect. Finally, greater stroke severity is associated with higher risk of neglect[5], and the severity of both the lesion itself and symptoms of neglect have previously been shown to predict poorer recovery[18,60]. Accumulating evidence suggests that neglect follows principles of proportional recovery[10,61,62], such that patients with severe symptoms show quantitatively greater improvements over time but are less likely to meet formal recovery criteria. Due to limited information on either the severity of stroke or neglect across studies, we could not formally evaluate this possibility. Of the studies reporting high stroke or neglect severity rates, two observed below-average recovery rates (<15%)[30,38], whereas one study found a 75% recovery rate[35]. Additionally, two studies showed that baseline symptoms were more severe in patients with chronic neglect compared with patients who recovered[36,41]. These findings suggest that acute severity could predict recovery rates. However, there is emerging evidence that the impact of severity on neglect recovery is complex and may vary depending on different factors, such as age[62] and neglect subtype[10]. Future studies exploring predictors of recovery should therefore consider potential interactions of severity and patient characteristics.

### Strengths and limitations

This review followed PRISMA guidelines to produce a methodologically robust synthesis of the literature on visuospatial neglect recovery. High levels of heterogeneity were observed across studies, limiting the precision of estimated recovery rates. We addressed this by carrying out sensitivity analyses which excluded low-quality studies, with resulting recovery rates being similar to the main meta-analyses. We also examined the contribution of patient and study characteristics to variability in meta-regressions. However, we acknowledge that our exploration of sources of heterogeneity was limited by the availability of primary data and could therefore not evaluate the impact of variables such as neglect subtype or severity. In addition, there was insufficient data to determine the influence of medical treatments or standard rehabilitation procedures on neglect recovery. Whilst thrombolysis and thrombectomy are known to improve outcomes after stroke [63,64], there were insufficient studies (n = 3) reporting administration rates to analyse the impact of these treatments on neglect recovery. Similarly, only five studies reported details on standard rehabilitation protocols for stroke survivors, such as provision of physiotherapy and occupational therapy.

The present review focused on spatial deficits which are central to the neglect syndrome. However, it is important to note that non-spatial impairments may exacerbate difficulties experienced by patients with neglect [65]. As the majority of studies did not use non-spatial measures of cognition, it remains unclear whether the presence of non-spatial cognitive impairments contributes to differences in recovery outcomes. Finally, included studies used a wide range of instruments for diagnosing neglect, which could lead to differences in sensitivity to neglect symptoms. However, descriptive data from a subset of studies using the BIT, a well-validated and standardised assessment for neglect, showed a pattern of recovery which was highly comparable to the main results.

### Clinical implications

Meta-analyses findings indicate that a large proportion of patients with neglect recover within the first six months after stroke. These findings provide useful information for both stroke survivors and clinicians, who benefit from having a clearer understanding of the prognosis for neglect and associated support needs. Additionally, estimates of neglect recovery can be applied to inform policy and care services as to the predicted needs of stroke survivors over time. It is important to emphasise that, whilst the present results suggest most stroke survivors will recover from neglect, approximately 40% with neglect are expected to have persistent impairments. Given that neglect is associated with poor functional outcomes and reduced rehabilitation efficacy, it is critical that clinicians are aware that patients with neglect may need continued support to cope with their symptoms. Whilst neglect severity appears to be a potential factor [10], further research is needed to converge on predictors of chronic neglect and to devise appropriate support and interventions for these patients.

## Conclusion

This systematic review and meta-analysis shows that 53% of stroke survivors with neglect recover within the first 6 months, with most natural recovery taking place within 3 months post-stroke. However, heterogeneity within the existent literature is high; further large-scale studies are needed to confirm the factors which influence neglect recovery, most likely lesion site and severity. Future studies on visuospatial neglect should prioritise the identification of predictors for chronic symptoms, as well as the development of support and interventions for stroke survivors with persistent neglect.

## Non-standard Abbreviations and Acronyms

BIT: Behavioural Inattention Task
JBI: Joanna Briggs Institute

## ACKNOWLEDGEMENTS

None.

