## Supplemental Materials for "The natural recovery of visuospatial neglect: a systematic review and meta-analysis"

### Supplemental Material

#### *Search strategy*

The following search was carried out on OVID to search the PsycInfo, Embase, and MEDLINE databases:

(stroke* or cerebrovascular accident* or ischemic stroke* or hemorrhagic stroke* or embolic stroke* or brain stem infarct* or thrombotic stroke* or paralytic stroke* or lacunar stroke* or completed stroke* or cardioembolic stroke* or syndrome of little stroke*, neurological or cerebellar stroke* or progression stroke* or occlusive stroke* or thromboembolic stroke* or stroke test response* or infarct* or cerebrovascular disease* or acute stroke* or CVA* or cerebrovascular apoplexy or vascular accident*, brain* or cerebrovascular stroke*).mp
AND
(neglect* or hemi#neglect* or sensory neglect* or visual neglect* or visuospatial neglect* or visual#spatial neglect or hemi#sensory neglect* or spatial neglect* or neglect of left side of body or hemi#motor neglect* or decreased spatial neglect* or unilateral neglect* or unilateral spatial neglect or hemi#sensory neglect or inattenti* or motor neglect* or vis* spatial percept*).mp
AND
((cogniti* assessment* or cogniti* test or cogniti* task or neuropsychology* test* or neuropsychology* assessment* or neuropsychology* task* or cancellation task* or target cancel* or copy*) and draw* task*) or copy* or clock test* or line bisect* or visual field task* or Catherine Bergego Scale or CBS or stroke impact scale or SIS or national institutes of health stroke scale or NIHSS or stroke impact scale or SIS or behavioural inattention test or BIT or oxford cognitive screen or OCS or Semi-structured Scale for Functional Evaluation of Hemi-Inattention or measure o measur*).mp

#### *Sensitivity analyses*

Sensitivity analyses excluded all studies rates as low quality on the JBI (n = 6). Meta-analyses indicated a recovery rate of 38% (CI = 20-58%, $I^2$ = 88%) in the early phase (0-3 months), based on 10 studies with 242 stroke survivors. Recovery rates increased to 53% (CI = 32-72%, $I^2$ = 91%) at 6 months post-stroke across 9 studies of 395 patients. There were no further changes in the late recovery phase (>6 months), with an estimated pooled prevalence of 56% (CI = 41-71%, $I^2$ = 78%) following analyses of 9 studies with 213 stroke survivors.

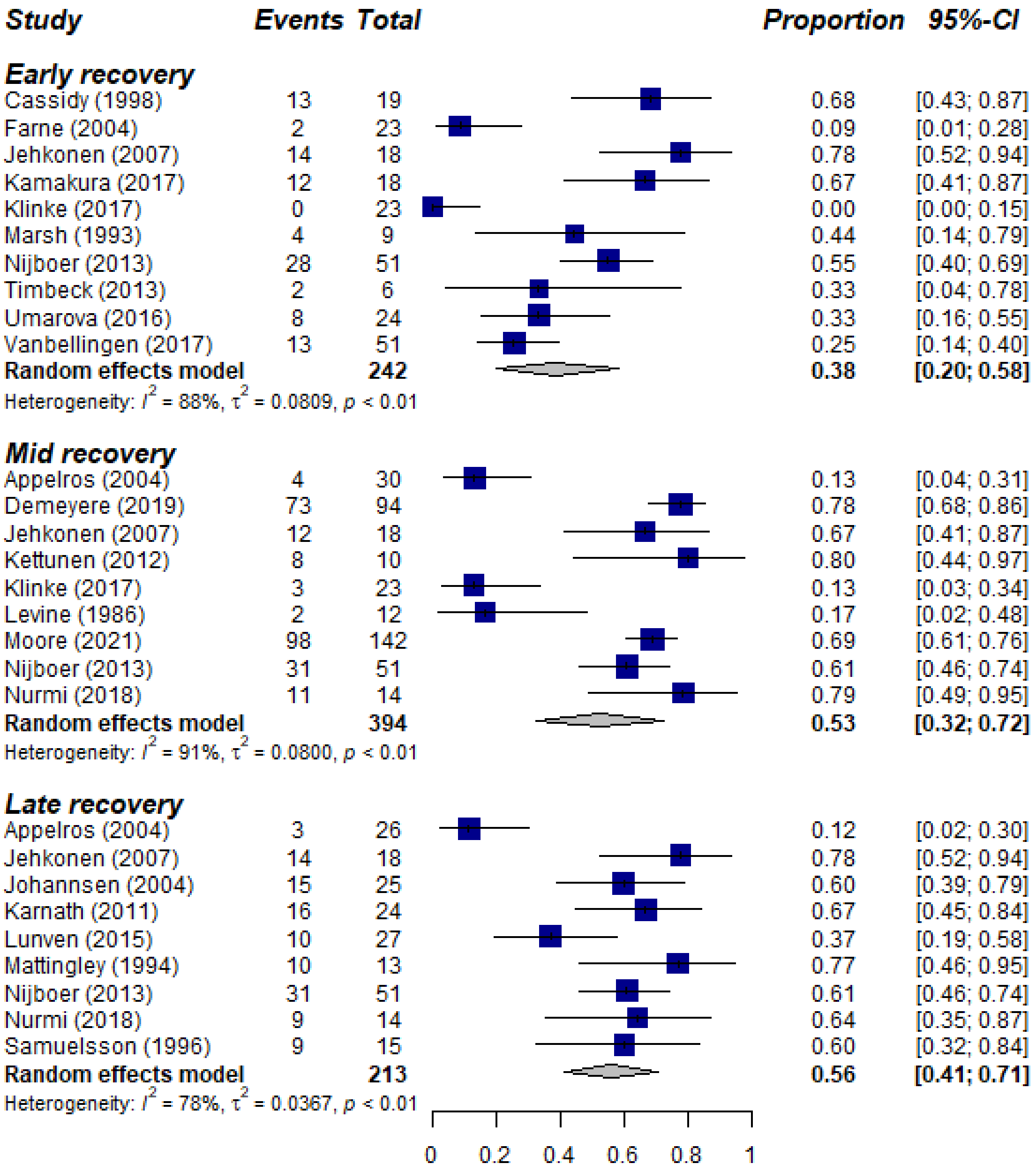


**Figure S1.** Forest plots of neglect recovery stratified by phase for studies of moderate and high quality
